# Could the load of carbapenemase genes in hospital wastewater be a proxy for the epidemiology of emerging resistance to carbapenems in humans?

**DOI:** 10.1101/2025.09.01.25334728

**Authors:** Camille Favier, Mylène Toubiana, Isabelle Zorgniotti, Olivier Courot, Franz Durandet, Patricia Licznar-Fajardo, Estelle Jumas-Bilak

## Abstract

**Background:** Antimicrobial resistance (AMR) poses a growing threat to global public health and is a key concern for infection control teams in hospitals. However, AMR surveillance is time-consuming and limited in most countries, resulting in incomplete findings. In several high-income countries, infection control teams ensure the contact tracing of every patient carrying an emerging extremely resistant bacterium which is very time-consuming. Wastewater surveillance (WWS) has been proposed as an alternative approach for the surveillance of infectious diseases. This study aims to investigate the dynamics of endemic (*bla*_CTX-M_) and emerging AMR genes (*bla*_OXA-48_, *bla*_NDM_, *bla*_KPC_ and *vanA*) in wastewater under real-world hospital conditions, and to compare results from two hospital buildings with contrasting resistance epidemiology, haematology building versus traumatology / orthopaedic building.

**Methods:** The sampling programmes were adapted according to the sampling sites and patient flow for each hospital building. Genes were quantified in the effluent using qPCR and dPCR. Cultivable carbapenemase-producing Gram-negative bacteria were characterised using MALDI-TOF MS and PCR.

**Results:** The feasibility of AMR monitoring in wastewater in real hospital conditions was demonstrated by dPCR and qPCR, which produced correlated results. The presence of peaks and the low load of the *vanA* and *bla*_NDM_ genes in wastewater (compared to *bla*_CTX-M_) were consistent with their known emerging status, as indicated by national and local epidemiological data. However, the constant presence of *bla*_OXA-48_ and *bla*_KPC_ at levels often higher than those of *bla*_CTX-M_ in wastewater did not reflect the known epidemiology of these emerging resistances, particularly in the case of *bla*_KPC_. Bacterial culture also revealed discrepancies with known epidemiology, with a majority of *Citrobacter* spp. carrying *bla*_KPC_ and *bla*_OXA-48_ in wastewater, whereas *Escherichia coli* and *bla*_OXA-48_ dominated in patients. Quantifying carbapenemase genes in wastewater was able to differentiate between buildings housing patients with a high or standard risk of emerging AMR.

**Conclusion:** Alongside the encouraging findings regarding patient populations and the potential predictive power of AMR WWS, this study identified obstacles that need to be overcome before it can be used for routine surveillance in an infection control hospital context.

## Introduction

Wastewater is a complex matrix that transports a wide range of chemical and biological molecules. The analysis of drug residues in wastewater to monitor public health coined the concept of wastewater-based epidemiology (WBE), also called wastewater surveillance (WWS) [Vitale et al., 2021]. After several attempts at WWS for viral infections [Benschop et al., 2021], the monitoring of SARS-CoV-2 in wastewater as a proxy for human epidemiology highlighted the utility of WWS for emerging infectious diseases [3]. WWS is simpler and less costly than individual testing of a human population, particularly for infectious agents that are often associated with asymptomatic carriage, such as SARS-CoV-2 [Polo et al., 2020] and multi- or extremely drug-resistant bacteria [Pruden et al., 2021].

Antimicrobial resistance (AMR) is a growing threat to global public health because it can lead to increased morbidity and mortality from treatment failures, and can also slow progress in critical care or surgery [Naghavi et al., 2020]. The emergence, spread and increasing prevalence of carbapenemase-producing *Enterobacterales* (CPE) is of particular concern because carbapenemases inactivate carbapenems, which are considered the antibiotics of last resort for serious Gram-negative infections. As a result, CPE has been identified by the World Health Organization (WHO) as one of the top three critical priorities [WHO, 2024]. CPE was spreading in an epidemic mode, while other multidrug-resistant bacteria also identified by the WHO as critical priorities, such as extended-spectrum beta-lactamase (ESBL)-producing bacteria (mainly CTX-M-producing bacteria), are now spreading in an endemic mode [Castanheira et al., 2021]. In France, the most common gene associated with CPE is *bla*_OXA-48_, followed by *bla*_NDM_ and less common genes (*bla*_KPC_, *bla*_VIM_, *bla*_IMI_, *bla*_GES_…) [CNR, 2022]. In addition to CPE, vancomycin-resistant *enterococci* (VRE) are emerging as a major cause of healthcare-associated infections and are among the highest global public health priorities [WHO, 2024]. VRE express the *vanA* or *vanB* genes, which cause cell wall modifications and confer resistance to vancomycin and other glycopeptides that are antibiotics of last resort.

In most European countries, surveillance for CPE and VRE is based on voluntary reporting by medical laboratories or hospital infection control teams (ICTs). In the hospital, detection of asymptomatic carriage is achieved by individual rectal screening, either systematically for patients in intensive care units or targeted only at patients at high risk of carriage, such as those hospitalised in high endemic areas or who have been in contact with a CPE or VRE-carrying patient. Contact tracing for each CPE or VRE case, as well as individual screening of high-risk patients, is poorly received by patients. It is also burdensome and time-consuming for healthcare workers, and it only provides patchy epidemiological data. There is clearly a need for more efficient tools to facilitate patient follow-up and enable more comprehensive surveillance of critical emerging resistant pathogens. Given the faecal shedding of CPE and VRE, WWS of emerging AMR may be a reasonable option. The post-COVID-19 surge in WWS indications, combined with strong political will for the development of environmental AMR surveillance, could rapidly lead to the establishment of regulatory obligations for AMR WWS [Hutinel et al., 2021]. However, the correlation between WWS and classical surveillance, i.e. between the antimicrobial resistance gene (ARG) signal in wastewater and the frequency of isolation of a particular resistant bacterium in medical laboratories, remains rarely evaluated [Hutinel et al., 2021; Flach et al., 2021, Urase et al., 2022].

Here, we used qPCR and dPCR to quantify the *bla*_CTX-M_, *bla*_OXA-48_, *bla*_NDM_, *bla*_KPC_ and *vanA* genes in hospital wastewater. The aim was to compare the dynamics of endemic and emerging AMR genes in France and to compare the data from two hospital buildings that host patients with contrasting resistance epidemiology. Finally, the study highlights the value of AMR WWS but also some limitations that must be considered before implementing it in the routine of infection control.

## Material and methods

### Hospital wastewater sampling

The University Hospital of Montpellier, in the south of France, is a 2600-bed tertiary care teaching hospital. Hospital wastewater sampling was carried out using automatic samplers supplied by the IAGE Company (Montpellier, France) in distinct buildings with independent wastewater collection pipes and tanks. The sampling and sub-samples storage were done from 12 to 15 °C along the sampling period. The GPS coordinates of the sampling sites in the Montpellier University Hospital were as follows: 43.630501792764484, 3.850851011755367 (SRR site) and 43.63036309242587, 3.864716021935508 (HRR site). Samples were conducted between 05-11-2022 and 06-02-2022 in SSR building (9 samples), and between 03-07-2022 and 05-23-2022 in HRR building (12 samples).

The sampling programmes were designed to obtain the theoretical volume of 1 L. In the SRR building, 24-hours composite samples were collected in a wastewater retention tank at a sampling frequency of 20 seconds per hour. Sampling was continuous throughout the day with a break between 11:00 AM and 01:00 PM. In the HRR building, 1 L of wastewater was collected weekly in the main building drain, where the wastewater does not stagnate due to rapid flow with each use of the toilet, sink or shower. The frequency and volume of sampling was optimised according to the flow in the collector during the hours of maximum use of the toilets and bathrooms by patients: 1 minute of sampling every 2.5 minutes between 7.30am and 11am, then between 4.30pm and 8pm. Samples were transported in coolers, carefully homogenised and separated into two subsamples, one for qPCR and the other for dPCR.

### Total DNA extractions

For qPCR analyses, the MasterPure Gram positive DNA purification kit (Lucigen, LGC Biosearch Technologies) was used according to the manufacturer’s instructions (DNA extraction triplicates). Purifications were performed on the pellet obtained from 3 mL of wastewater (3 min centrifugation at 12,000 g). Quantities and qualities of DNA extracts were measured using a NanoDrop spectrophotometer (ThermoFisher Scientific). For dPCR, 30 mL of the homogenised wastewater were vortexed 3 times for 10 seconds each. The sample was cooled on ice for 15 seconds between each vortex burst. An Amicon Ultra-15 centrifugal filter unit (Merck Millipore, cut-off: 10 kDa) was hydrated with 2.5 mL of MQ water and spun at 3234 g for 10 minutes at 4 °C. Fifteen mL of the vortexed samples were transferred in the Amicon Ultra-15 Centrifugal Filter Unit and were then concentrated by centrifugation at 3234 g for 35 min at 4 °C (ultrafiltration). The extraction of total DNA was performed using the IndiMag Pathogen Kit (Indical Bioscience) on the total volume of the concentrated sample corresponding to 15 mL of wastewater. DNA extracts from both methods were then stored at −20 °C until use.

### Genes quantification by qPCR and dPCR

qPCR reactions were performed using SYBRgreen chemistry on a LightCycler Nano (Roche) in eight-well strips for the quantification of six target genes: 16S rRNA gene [Maeda et al., 2003], *bla*_CTX-M_ [Marti et al., 2013, Lartigue et al., 2007], *bla*_OXA-48_, *bla*_NDM_, *bla*_KPC_ [Subirats et al., 2017, Poirel et al., 2011] and *vanA* [Mirzae et al., 2013]. Each reaction contained SensiFAST SYBR No-ROX mix 1X (Bioline Meridian Bioscience), 0.4 µM of forward and reverse primers (Supplementary Table S1), 1 µL of 10-fold diluted DNA (10, 100 and 1000-fold diluted DNA for 16S rRNA gene amplification) and sterile water to a final volume of 10 µL.

**Table 1.** Epidemiological data on the resistance markers targeted in this study (France, 2022)

|  | CPE |  |  | ESBL-E | VRE |  | Data sources |
| --- | --- | --- | --- | --- | --- | --- | --- |
| Percent of resistant isolates among total tested isolates, nationwide | 0.1-1*% |  |  | 8-25*% | 0.7% |  | [ECDC] |
| Percent of gene types, nationwide (percent of gene types, in the hospital) | *bla*<sub>OXA-48</sub><br>60,6%<br>(74%) | *bla*<sub>KPC</sub><br>2,1%<br>(0%) | *bla*<sub>NDM</sub><br>25,7%<br>(26%) | *bla*<sub>CTX-M-15</sub><br>74-92*%<br>(na) | *vanA*<br>82,6%<br>(100%) | *vanB*<br>14,8%<br>(0%) | [CNR, 2023]; ICT unpublished data |

Values in parentheses corresponds to the hospital data provided by ICT for this study. na for not available. \*Depending on the species. CPE for Carbapenemase-Producing *Enterobacteria*; ESBL-E Extended-Spectrum Beta-lactamase-producing enterobacteria, VRE for Vancomycine Resistant *Enterocci*.

qPCR cycling conditions started with polymerase activation at 95 °C for 3 min, followed by 40 cycles composed by 10 s at 95 °C, 10 s at 60 °C (16S rRNA gene, *bla*_NDM_, *bla*_OXA-48_), at 62 °C (*bla*_CTX-M_ and *bla*_KPC_) or at 58 °C (*vanA*), and 10 s at 72 °C. Finally, amplification products were gradually heated from 60 to 95 °C (0.1 °C/s) in order to obtain the melting temperatures and to check specificity. Standard curves were obtained by quantification of 10-fold serial dilution of known quantities of linearized plasmids containing specific amplicons. DNA extracted from in lab collection strains carrying *bla*_OXA-48_ (*Klebsiella pneumoniae* ARS619), *bla*_NDM_ (*Escherichia coli* B26), *bla*_KPC_ (*E. coli* ARS224), *bla*_CTX-M_ (*K. pneumoniae* 2M2E5) and *vanA* (*Enterococcus faecium* strain T301-1) genes were used as positive controls, and sterile water as negative control. For each sample, quantification was performed in technical duplicate. For inhibition assays, i) the same protocol was used to quantify the *bla*_NDM_ gene in a reaction containing a known amount of standard plasmid in addition to 1 µL of 10-fold diluted DNA extracted from a wastewater sample known to be negative for this gene and ii) quantifications of 16S rRNA genes from 10, 100 and 1000-fold diluted DNA extracts were compared. qPCR quantification data were only considered if the amplification had a specific Tm, if the quantification was above the limit of quantification (Supplementary Table S2) and if it was i) present in both technical replicates and ii) present in at least two of the three DNA extraction replicates. If this was not the case, the gene was considered to be detected but not quantifiable. Data were analysed using the LightCycler Nano Software suite (v1.0).

dPCR reactions were conducted using a 5-plex assay developed by IAGE on a QIAcuity Eight Plateform System, using the QIAcuity Probe PCR Kit and QIAcuity Nanoplates 26 K 24-wells (Qiagen, Germany). The same primers as for qPCR assays were used, supplemented with hydrolysis probe (Supplementary Table S1). The dPCR reaction mixtures were prepared in a standard PCR plate as follows: each reaction contained 10 µL of 4X Probe PCR Master Mix, 5 μL of the primers/probe mix (2.25 µM of both primers, 0.625 µM of probe), 4 µL of DNA extract and nuclease free water to a final volume of 40 µL. The reaction mixtures were transferred into a QIAcuity Nanoplate and the plate was loaded onto the QIAcuity Eight automated system. The workflow included (i) a priming and rolling step to generate and isolate the chamber partitions (26,000 partitions), (ii) an amplification step with the following cycling protocol: 95 °C for 2 min for enzyme activation, 95 °C for 5 s for denaturation, and 58 °C for 60 s for annealing/extension for 40 cycles; and (iii) imaging step by reading fluorescence emission after excitation of the probe at the appropriate wavelength. Data were analysed using the QIAcuity Software suite v3.1.0.0.

qPCR and dPCR results obtained in gene copy number per µL of DNA were both transformed in copy number mL-1 of wastewater. Normalised genes abundances were calculated as the ratio between resistance genes and the 16S rRNA gene copy numbers. These data were plotted on a logarithmic scale (log10). The gene *bla*_KPC_ was not quantified by dPCR but by qPCR only.

### CPE cultures and species identification

Fifty µL of pure, 10-fold and 100-fold diluted samples were plated on CHROMagar mSuperCARBA (Graso Biotech) medium using an easySpiral automatic plater (Interscience) in exponential and constant modes, allowing colorimetric presumptive identification of *Enterobacterales*. After overnight incubation at 37°C, colonies were counted by morphotype and colour, and selected for subculture on the same medium. After an overnight incubation at 37 °C, each selected strain was frozen at −80 °C in trypticase soy broth (DIFCO) supplemented with 20% glycerol, which allowed the constitution of a library of 145 bacterial strains. Bacterial identification was performed on overnight cultures on trypticase soy agar using matrix-assisted laser desorption ionization time-of-flight mass spectrometry (MALDI-TOF MS) (Brucker). The threshold for species affiliation was 2.0 according to the in vitro diagnosis protocols of the supplier (Brucker).

### End-point PCR of *fimH*, *bla*_NDM_, *bla*_OXA-48_, *bla*_KPC_ and *bla*_CTX-M_ genes

Endpoint PCR amplifications for resistance genes were performed with the primer indicated in Supplementary Table S1. The *fimH* gene was amplified according to Johnson et al. (2000), (Supplementary Table S1). PCR amplifications were done on a Hybaid® thermocycler, starting with an initial denaturation for 2 min at 95 °C, followed by 35 cycles consisting in 30 s (45 s for *fimH*) at 95 °C, 30 s (45 s for *fimH*) at 60 °C (*bla*_NDM_, *bla*_OXA-48_), 62 °C (*bla*_CTX-M_), 55 °C (*bla*_KPC_), or 58 °C (*fimH*), 30 s (60 s for *fimH*) at 72 °C and a final extension for 7 min at 72 °C. Reactions contained GoTaq Flexi Buffer 1X and 0.625U of GoTaq DNA Polymerase (Promega), 1.75 mM of MgCl2, 0.2 µM of each primer (Supplementary Table S1), 25 ng of DNA from wastewater (for *fimH*) or 1 µL of boiling/freezing crude extract from bacterial strains for the other genes. Sterile water was used as negative control. Positive controls were DNAs extracted from a laboratory collection of bacterial strains: *E. coli* T352 for *fimH* gene and *K. pneumoniae* 21DU, ARS619, Fonc147, 2FA2E1 for *bla*_NDM_, *bla*_OXA-48_, *bla*_KPC_ and *bla*_CTX-M_ genes, respectively. Endpoint PCR results were visualised on 1.5 % agarose gel with ethidium bromide after electrophoresis with TriDye 1 kb DNA Ladder (New England Biolabs).

### Epidemiological and clinical data

The Infection Control Team (ICT) data used in this paper are Excel files that collate the species and resistotypes of all bacteria isolated from hospital patients. These data are generated by the medical microbiology laboratory and exported to the ICT surveillance file. Regional and national data about CPE and VRE were obtained from the reference national centre for antimicrobial resistance [CNR, 2023] and from the European Centre for Disease Prevention and Control [ECDC].

### Statistical analysis

A Shapiro-Wilk test was used to determine whether the variables followed a Laplace-Gauss normal distribution. To study the correlations between dPCR and qPCR data, and between absolute and normalised quantifications, Pearson’s or Spearman’s correlation coefficients (r or rs) were calculated depending on whether the data followed a normal distribution, and the nullity of these coefficients were tested. Box-plots were performed to study the distribution of the data of the different sites of sampling and comparison between sites were made using a t-test for variables following a normal distribution, or a Mann-Whitney test for those that did not follow a normal distribution. Fisher’s exact test was used to compare the proportion of CPE and carbapenemase producing bacteria between the two buildings. Simpson’s Diversity Index (D) was calculated for the two buildings using the formula 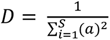 where (S) is the number of species and (a) is the proportional abundance of the nth species. Mann-Whitney test was conducted to compare the diversity indices between the two buildings. For all tests, rejection of the H0 hypothesis was considered significant when the p-value ≤ 0.05. All statistics were performed with GraphPad Prism software 8.0.1.

## Results

### Hospital buildings and patients’ populations in relation to CPE and VRE

The first hospital building has been considered as a standard resistance risk building (SRR) because the incidence of multidrug resistant bacteria was 0.44/1000 hospitalisation days in 2022. The patients cared daily in SRR for short-stays mainly for orthopaedic diseases (median length of stay about 4.5 days in 450 beds). During the sampling period of 4 weeks, approximately 7200 patients contributed to the wastewater of this building. Most of the SRR patients were not screened for resistant bacteria unless they had a known individual risk, such as a recent hospitalisation in a high-risk country or a history of contact with a patient carrying CPE or VRE.

On the opposite, the second building hosted patients with high risk of resistance (HRR) due to the general use of broad-spectrum antibiotics for proliferative blood disorders associated to immunosuppression (median length of stay about 10 days in 60 beds). The incidence of multidrug resistant bacteria in HRR was 9.39/1000 hospitalisation days in 2022. HRR patients were screened weekly for resistant bacteria. To ensure that most of the patient population changed between each sample, the wastewater from the HRR was analysed once a week for 11 weeks (1120 patients, from 07 March to 23 May 2022). No CPE or VRE outbreak was declared in SRR or HRR during the study period. The difference between the two building for patients AMR risks was confirmed by the difference in presence of patients who carried CPE or VRE during the study period (**Figure 1**).

**Figure 1.**
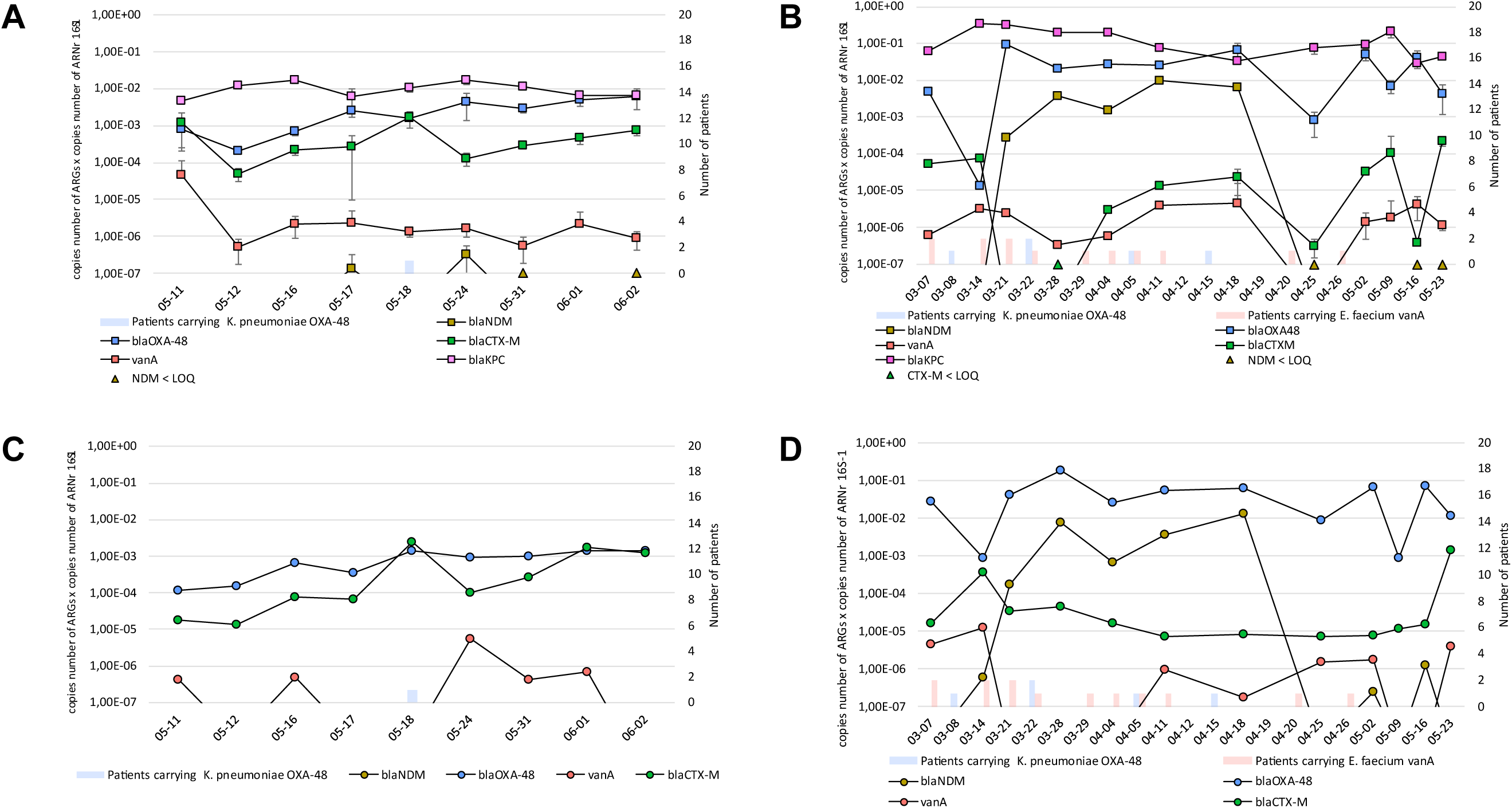
Normalised quantifications of resistance genes in wastewater of two hospital buildings. The genes *bla*_CTX-M_, *bla*_OXA-48_, *bla*_KPC_, *bla*_NDM_ and *vanA* were quantified using qPCR in wastewater of (A) standard resistance risk (SRR) and (B) high resistance risk (HRR) buildings, and using dPCR in wastewater of (C) SRR and of (D) HRR buildings. Quantifications are the mean of DNA extraction triplicates. An upward-pointing triangle on the x-axis indicates that the gene was detected, but its quantity was below the quantification limit in qPCR. The dPCR quantification of *bla*_KPC_ was not performed. The number of patients detected as carriers of CPE and VRE during the sampling period are indicated coloured bars in secondary y axis.

### Comparison of two protocols for quantifications of ARG in wastewater

The complexity of the wastewater matrix favours PCR inhibition. Comparative quantification of *bla*_NDM_ in a reaction containing a known amount of standard (*bla*_NDM_ plasmid) with or without diluted DNA extracted from wastewater negative for *bla*_NDM_ showed no significant difference (440 ± 98.12 copies mL-1 and 487 ± 13.49 copies mL-1, respectively). The absence of PCR inhibition was also confirmed for each sample by the results of 10-, 100- and 1000-fold diluted DNA 16S rRNA gene quantification (**Supplementary Figure S1**). The normalised abundances of AMR genes were positively correlated with the absolute abundances of AMR genes by both qPCR and dPCR (rsqPCR = 0.5 to 0.9, p-value ≤0.01to ≤ 0.0001; rsdPCR = 0.33 to 0.91, p-value ≤0.01 to ≤ 0.0001, except for *bla*_OXA-48_ (p-value = 0.2)).

**Figure 1** shows the qPCR and dPCR quantifications normalised for the 21 wastewater samples. Most of the 21 samples from the 2 hospital buildings gave qPCR and dPCR signals. Most of the unquantifiable positive signals in qPCR (7 points out of 21) corresponded to negative signals in dPCR suggesting either non-specific signal in qPCR or slightly better sensitivity of qPCR. In few cases, a qPCR signal higher than dPCR signal for about 1 log10 was observed, for instance for *bla*_OXA-48_ and *bla*_CTX-M_ in HRR samples (**Figure 1**). However, the results of Spearman’s correlation tests showed that there was a significant positive correlation between dPCR and qPCR normalised quantifications for *bla*_CTX-M_, *bla*_OXA-48_, *bla*_NDM_ and *vanA* genes (rs = 0.5 to 0.8, p-values ≤0.01 to ≤0.001). Since dPCR is considered to be unaffected by PCR inhibitors, the DNA extraction and qPCR protocols were again validated. This demonstrated that both methods are suitable for use in hospital settings for WWS, with no significant discrepancies.

### Links between resistance genes in hospital wastewater, patients carrying these genes and AMR epidemiology

**Table 1** presents nationwide epidemiological data on emerging extensively drug-resistant bacteria (CPE and VRE), alongside their associated resistance genes. The low percentage of resistant isolates (< or = 1%) confirms the emerging nature of CPE and VRE in France.

By contrast, (ESBL-E) are multidrug-resistant bacteria representing 8 to 25% of the enterobacterial isolates, depending on the species. This high prevalence rate suggests endemic diffusion in France. In this study, the beta-lactamase gene the most frequently associated with ESBL-E, *bla*_CTX-M-15_, has been used as an indicator of the endemic diffusion of antimicrobial resistance.

As expected, the ESBL marker *bla*_CTX-M_ was detected in all wastewater samples in both SRR and HRR buildings (**Figure 1**). The overall kinetics of *bla*_CTX-M_ and *bla*_OXA-48_ were very similar in SRR whereas *bla*_OXA-48_ is quantified at a higher level (1 log higher for most samples, p-value <0.0001) than *bla*_CTX-M_ in HRR (**Figure 1**). As *bla*_OXA-48_ is still emerging in France and in the hospital, the constant detection of *bla*_OXA-48_ in both SRR and HRR, sometimes at higher levels than *bla*_CTX-M_ was unexpected. Only 36 patients carrying OXA-48 CPE were identified in the entire hospital in 2022 (ICT surveillance data). During the study period, only two outpatients carrying OXA-48 *K. pneumoniae* stayed in HRR for several very short stays from 22 February to 16 April 2022 (**Figures 1B and 1D**). The dynamics of *bla*_OXA-48_ in the HRR effluent could not be fully explained by the presence of these two outpatients, as the signal remained at the same level after their discharge from 26 April until the end of the study (23 may). The persistent and high load of *bla*_KPC_ in both SRR and HRR (**Figure 1**) is even more surprising given the low rate of KPC CPE in France and the absence of KPC CPE detected in our hospital in 2022 (**Table 1**). The *bla*_NDM_ gene was detected less frequently. It was quantified or detected below the limit of quantification in 14 of the 21 samples. In SRR, *bla*_NDM_ was quantified at low level in only two samples (**Figures 1A and 1C**) whereas in HRR we observed a sudden increase between 22 March and 18 April (**Figures 1B and 1D**). After this peak, the *bla*_NDM_ signal rapidly became negative or at the limit of detection until the end of the study. These dynamics reflect the emerging status of the NDM-carbapenemase in patient epidemiology in France and in the hospital (**Table 1**). The sudden increase in HRR suggested that a patient or a group of patients carrying NDM bacteria were cared in HRR wards. However, no patient carrying NDM CPE was detected by individual biological diagnosis during this period (ICT surveillance data).

The dynamics of *vanA* gene during the study is shown in **Figure 1**. In qPCR, it was detected in all but one sample (24 April in HRR) in both buildings. The *vanA* gene load is lower (1 to 3 log) than that of *bla*_CTX-M_, considered as the standard for endemic gene, in SSR (**Figures 1A and 1C**). This was also the case in HRR, but for this ward *vanA* exceeded *bla*_CTX-M_ in few samples (21 March, 28 March and 16 May) (**Figures 1B and 1D**) while *vanA* resistance was still considered emerging in France and no outbreak was detected in HRR. During the study period, a maximum of 2 HRR patients per day were known as carriers of E. faecium *vanA* (**Figures 1B and 1D**). From 7 March to 11 April, the effluent signal appeared to be partially linked to the number of known *vanA* carriers. After 12 April, similar *vanA* signals were observed, despite only one outpatient being admitted twice for one day in the ward (ICT surveillance data). In conclusion, *vanA* levels in hospital wastewater were partially correlated with the presence of individual patients who carried the gene. The *vanA* dynamics in wastewater of the two buildings suggested a low-level of endemicity for *vanA* resistance in hospital patients.

### Does resistance gene monitoring in hospital wastewater differenciate patients populations?

The two hospital buildings examined in this study serve different patient populations in terms of their risk of carrying CPE and VRE: high risk for HRR patients and standard risk for SRR patients. The difference in normalised quantification of *bla*_CTX-M_ was highly significant between SRR and HRR, but unexpectedly *bla*_CTX-M_ was more prevalent in SRR (**Figure 2A**).

**Figure 2.**
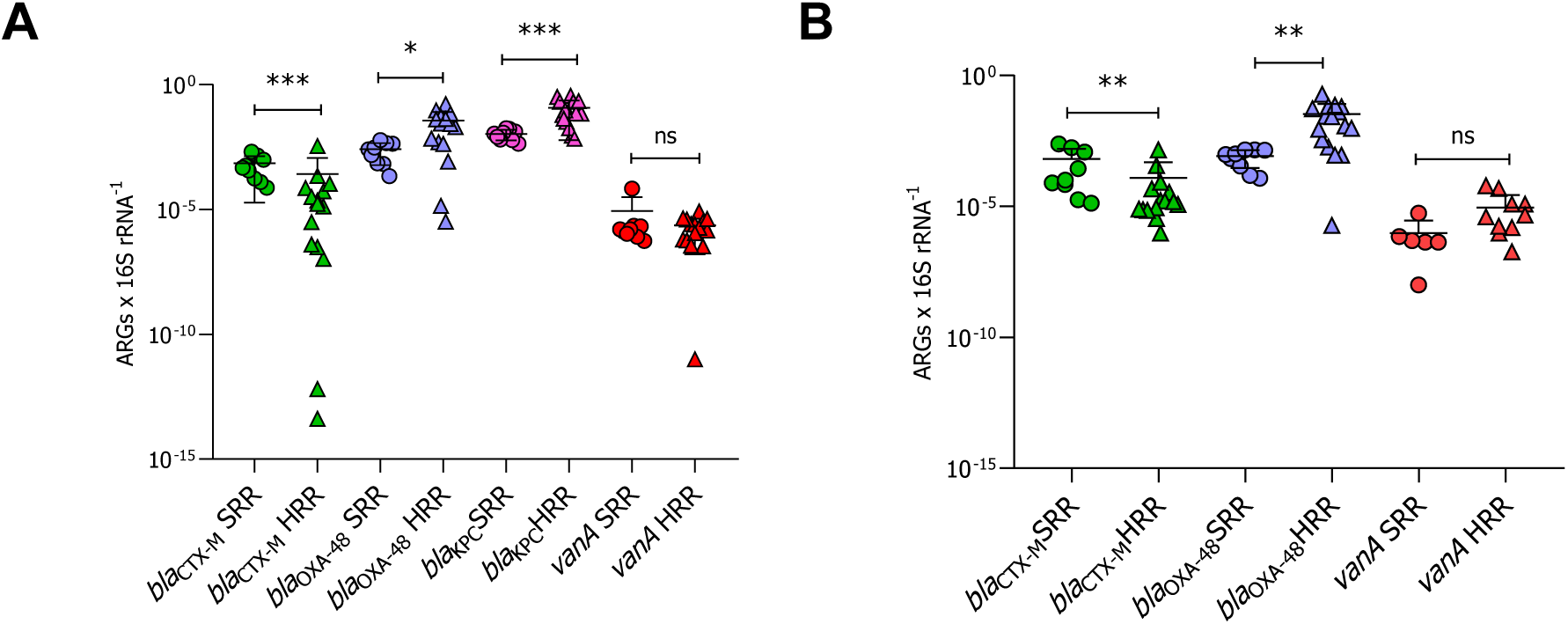
Distribution of the normalised loads of *bla*_CTX-M_, *bla*_OXA-48_, *bla*_KPC_ and *vanA* in wastewaters. (HRR), high risk resistance building and (SRR), standard risk resistance building. (A) quantification using qPCR; (B) quantification using dPCR (B). The results are expressed in an inverse logarithmic scale. Comparison of ARGs quantification between hospital buildings was performed by the Mann-Whitney test, and p-values are presented above the box plots (ns : non-significant; * : 0,01 < p-value ≤ 0,05; *** : 0,0001 < p-value ≤ 0,001; **** : p-value ≤ 0,0001). Median and IQR are presented on each scatter plot. The dPCR quantification of *bla*_KPC_ was not performed.

One explanatory hypothesis is antimicrobial stewardship in HRR wards, where any sign of systemic infection leads to the immediate prescription of broad-spectrum antibiotics that cover at least ESBL-E. This could limit the load of *bla*_CTX-M_ in patient microbiota and in HRR building effluent. The loads of carbapenemase genes, *bla*_OXA-48_ and *bla*_KPC_, were significantly lower in SRR wastewater than in HRR (**Figures 2A and 2B**). This was also the case for *bla*_NDM_, even if the high rate of negative signals prevented statistical analysis (**Figure 1**). These results suggested that the effluent loads of ESBL and carbapenemase genes in HRR and SRR are specific to the patient populations cared for in the respective buildings. However, the load of *vanA* was not significantly different between SRR than HRR (**Figures 2A and 2B**).

### Does the resistant culturable gammaproteobacterial community in wastewater reflected gene load in wastewater and local epidemiology ?

The loads of gamma-proteobacteria growing onto media selective for carbapenem resistance are showed in **Figure 3**. The bacterial load in the effluent varied from 104 to 2.108 CFU mL-1 in SRR and from 3.105 to 5.108 CFU mL-1 in HRR. The higher load of carbapenem-resistant bacteria in the HRR follows the trends observed for ARG load in the two buildings. The dynamics of resistant bacteria showed peaks unrelated to the summation of the normalised loads of the 3 carbapenemase genes (**Figure 3**) and the 16S rRNA gene dynamics (data not shown).

**Figure 3.**
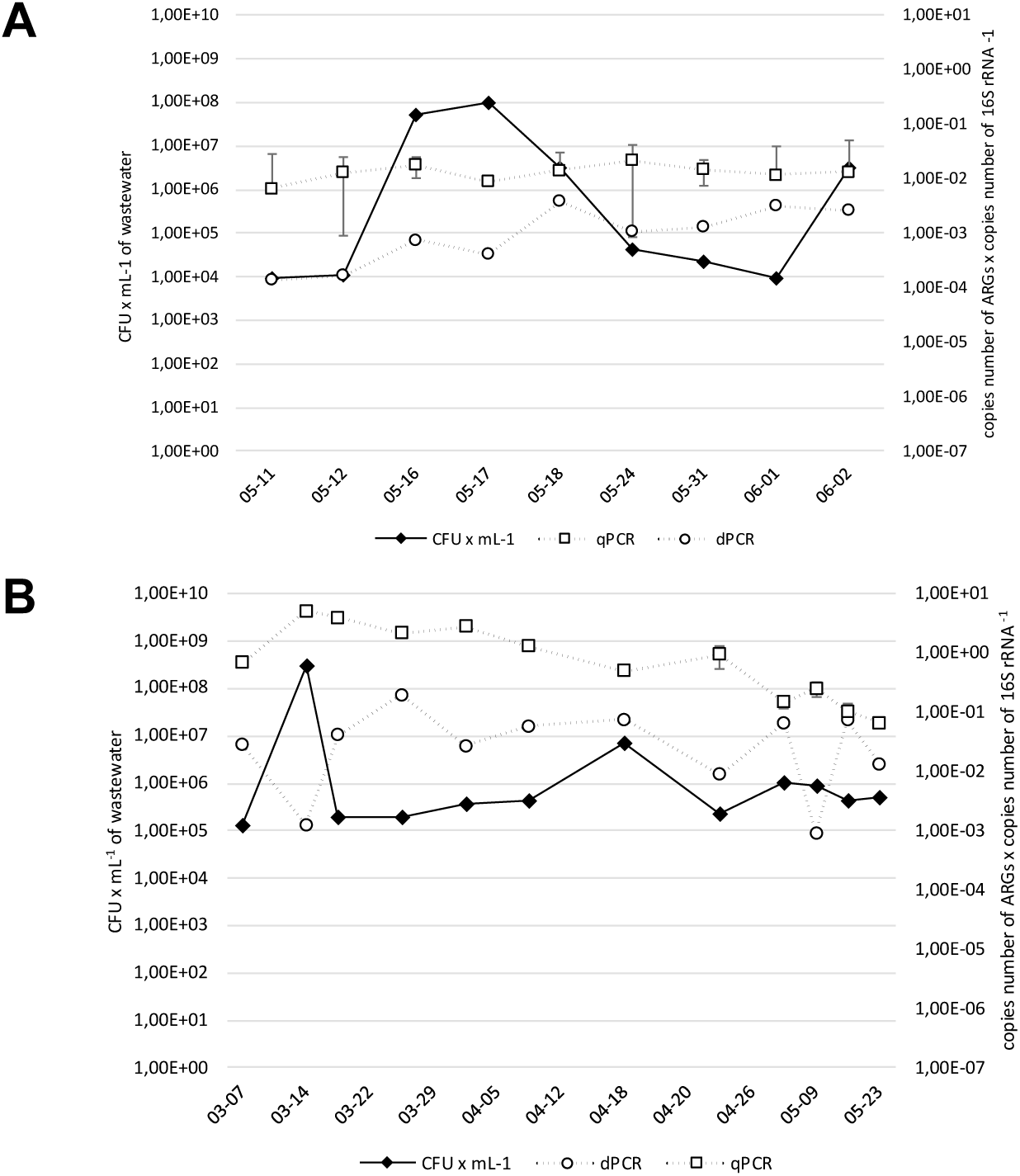
Load of carbapenemase resistant bacteria versus summation of normalised loads of carbapenemase genes. (A) standard resistance risk (SRR) building; (B) high resistance risk (HRR) buildings. Bacteria are grown onto mSuperCARBA™ medium. Carbapenemase genes are quantified by dPCR and qPCR. and high resistance risk (HRR) (B) buildings.

A total of 145 strains were selected on the basis of their colony morphotypes. Simpson’s Diversity Index (D) was calculated for HRR (0.67 ± 0.08) and SRR (0.72 ± 0.08) buildings. Mann-Whitney test performed to compare the diversity in the two buildings yielded a statistic W of 39 and a p-value of 0.20. The results indicate that there is no statistically significant difference between the taxonomic diversity of wastewater in the two buildings.

More than half of the selected bacteria (76/145 = 52%) resisted to carbapenems by carrying the carbapenemase encoding genes *bla*_OXA-48_, *bla*_KPC_ or *bla*_NDM_. Fourteen different species belonging to 7 genera of carbapenem-producing gamma-proteobacteria were identified including 4 genera of *Enterobacterales*: *Citrobacter, Enterobacter, Klebsiella and Kluyvera* (**Figure 4**).

**Figure 4.**
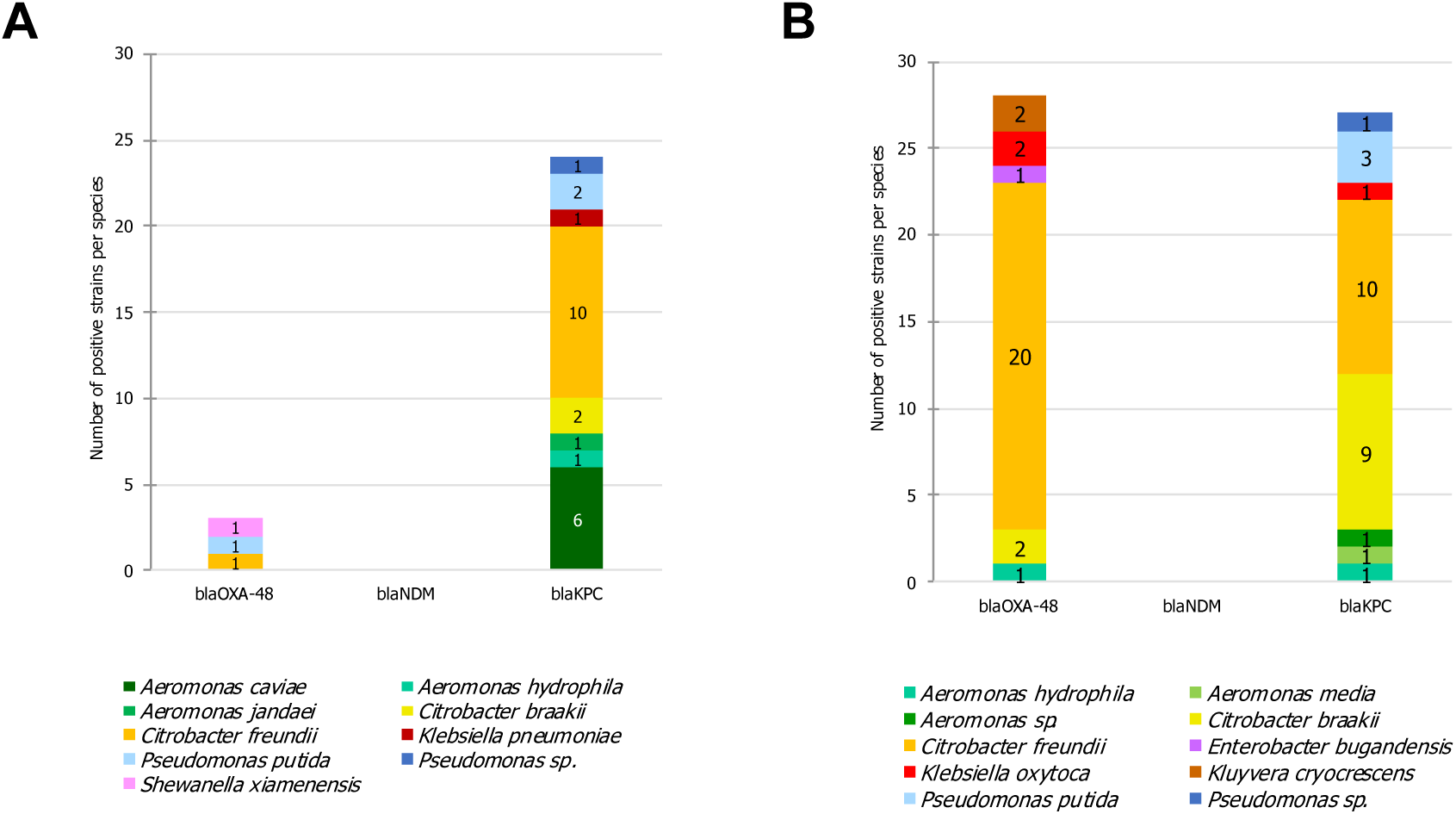
Identification and resistotype of carbapenemase-producing species. Bacteria isolated from SRR (A, n_tot_ = 81) and HRR (B, n_tot_ = 64) wastewaters onto mSuperCARBA™ medium.

The most common carbapenem resistotypes in *Enterobacterales* were OXA-48-producing *Citrobacter freundii* (n=20 in HRR and n=1 in SRR), KPC-producing *C. freundii* (n=10 in each building) and KPC-producing *Citrobacter braakii* (n=9 in HRR and n=2 in SRR). Six strains carried both *bla*_OXA-48_ and *bla*_KPC_ gene (4 *C. freundii*, 1 *C. braakii* and 1 *Klebsiella oxytoca*), all of them were isolated from HRR wastewater (**Figure 4**). Finally, forty-nine of the 53 Citrobacter spp. strains isolated in this study encoded *bla*_OXA-48_, *bla*_KPC_ or *bla*_NDM_. The rate of viable/cultivable OXA-48-, NDM- and KPC-CPE among strains isolated on selective medium was significantly higher in HRR (65%) than in SRR (41%) (Fisher’s exact test, p-value = 0.0042). Furthermore, the majority of bacteria growing on the carbapenem resistance selective medium resisted to carbapenem via OXA-48 and KPC-carbapenemase production in HRR, but not in SRR where KPC CPE largely dominated (**Figure 4**).

The pathogens *E. coli* and Pseudomonas aeruginosa was not isolated in this study. Concerning *E. coli*, the specific *fimH* gene was searched in wastewater and was detected in every wastewater sample (data not shown). This suggests that although *E. coli* DNA is present in wastewater, carbapenemase-producing *E. coli*, if present, are no longer viable or cultivable in the samples tested.

The ICT data from the routine surveillance of CPE in hospital showed that the most commonly isolated CPE from patients in the Montpellier hospital, all buildings combined, in the study period, were *K. pneumoniae*, *E. coli*, *Enterobacter cloacae* and *C. freundii* (**Figure 5**). This clearly differed from wastewater, where Citrobacter dominated and *E. coli* was not detected. **Figure 5** also showed that, with the exception of *bla*_OXA-48_, which is common in both human and wastewater, the proportion of bacterial species resistant to carbapenems by production of NDM and KPC differed between human and wastewater. In conclusion, the taxonomic diversity and resistotypes of CPE isolated from patients did not match those of resistant bacteria circulating in wastewater.

**Figure 5.**
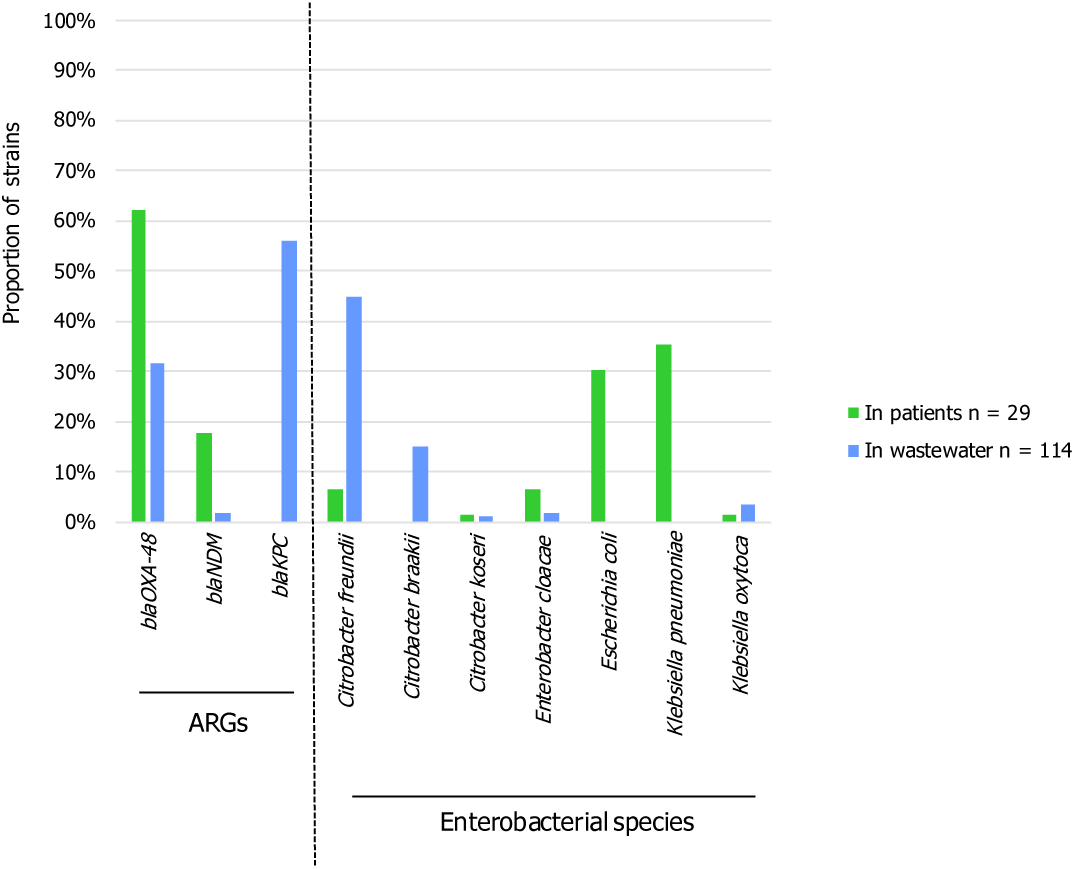
Distribution of resistotypes and taxa of carbapenemase-producing *enterobacteria* in patients and hospital wastewater.

## Discussion

A recent systematic review [Tiwari et al., 2024] showed that most published studies on ARGs in wastewater have focused on the risk of spread from hospitals or other AMR hotspots into the environment, and the subsequent risk to public health [Sambaza et al., 2023]. Few papers, however, have addressed the WWS of AMR. While previous publications have presented ARGs in wastewater as potential AMR indicators for surveillance purposes, the concept of WWS applied to AMR surveillance has only recently been highlighted in publications such as an editorial in Science [Aarestrup et al., 2020] and a commentary by Larsson et al. (2023) in Nature Reviews Microbiology [Larsson et al., 2023]. The scientific basis for WWS of AMR was a comprehensive set of experiments based on metagenomics. These experiments demonstrated the reliability of WWS of AMR at national [Su et al., 2017] and global [Munk et al., 2022] levels. Furthermore, correlations with socio-economic, health, and environmental factors have been demonstrated [Hendriksen et al., 2019]. Approaches targeting resistant bacteria or ARGs of interest have also been developed, showing the correlation between AMR in wastewater and AMR epidemiology [Pärnänen et al., 2019;, Blaak et al., 2021, Huijbers et al., 2020]. If global surveillance is needed to inform national and international agencies, local or territorial surveillance, for example at the city level [Josef et al., 2019] or within hospitals [10, 31], could provide insights into the extent of antibiotic use, the local mode of bacterial resistance spread (endemic or epidemic), and thus guide effective antibiotic stewardship and infection control interventions.

This study proposes to monitor emerging resistance genes in wastewater of a hospital under real-world conditions. Specifically, the study design does not involve any changes in the structure of the wastewater network or in patient monitoring. Thus, feasibility and barriers has been assessed in a real hospital context. We examined wastewater from two buildings displaying different effluent systems: a retention tank (SRR building) and a drainpipe directly connected to the patients’ sanitary system (HRR building). To limit biases linked to these structural differences, we collected pooled samples, rather than grab samples, because pooling has been shown to be the optimal method for obtaining the highest bacterial diversity in hospital drains [Huijbers et al., 2023]. The tank, located between the proximal SRR outfalls and the municipal collector, probably increased the homogeneity of the effluent, thereby reducing the number of sub-samples required. In HRR, the flow in the outfall pipe was intermittent, depending on the use of the sanitation systems. Given the small contributing population and the short distance between the toilet and the sampling point in HRR, it was determined that very small and frequent subsamples would be required, in line with previous studies [Huijbers et al., 2023; Paulshus et al., 2019]. Because the sampling and storage protocol has been shown to affect bacterial diversity [Huijbers et al., 2023], we compared taxonomic diversity indices in HRR and SRR wastewater. Statistically similar Simpson’s index suggested that there was no significant sampling bias in the study and indirectly validated the sampling protocols.

Previous studies have shown that the enrichment of resistant bacteria and the reduction of *E. coli* due to the biocidal selection of hospital wastewater has been observed in vitro at 20 °C [Huijbers et al., 2023].]. The unexpected absence of culturable carbapenem-resistant *E. coli* despite the presence of *fimH*, an *E. coli*-specific gene, in each sample suggests a potential biocidal effect. It is noteworthy that *E. coli* is the most common organism among CPE isolates from patients in France and in the Montpellier hospital. A publication describing WWS in a Swedish hospital [10] reported a high prevalence of carbapenemase-producing *E. coli*. This suggests the possibility of a depletion of carbapenemase-producing *E. coli* in the Montpellier hospital effluent, as evidenced by the findings of Urase et al. (2022) [11] who detected only six *E. coli* among 247 carbapenem-resistant bacteria isolated from wastewater in Japan.

The first objective of the study was to compare the load of AMR genes in wastewater with epidemiological data from classical AMR surveillance in hospitals and across the country. The inconstant detection of *bla*_NDM_ is consistent with the epidemiological data from clinical microbiology of NDM-CPE, which is emerging in France and Europe. For the *vanA* gene that still emerging in France, WWS suggested rather a low endemicity in the hospital. It is noteworthy that before the study, in 2023, a large *vanA* outbreak occurred in several wards of the hospital (ICT, unpublished), suggesting that WWS could be predictive of a risk of local outbreak. The *bla*_OXA-48_ gene is consistently detected, sometimes at a higher level than the *bla*_CTX-M_ gene, which is used as a reference for endemic resistance. This suggests a bias due to a resistant microbial community in the network or calls into question the current emerging status assigned to OXA-48 CPE in our hospital and in France. Current surveillance, which is only targeted at patients who are supposed to be at high risk of carriage, could overlook the rate of OXA-48 CPE in the general population. The dominance of *bla*_KPC_ in hospital effluents was the most surprising finding of this study, given that KPC-producing bacteria are in the minority in France [8]. Similarly, the wastewater-resistant bacterial community was clearly dominated by carbapenemase-producing Citrobacter spp., which are sporadically detected in patients in 2022 in France. At our hospital, the *bla*_KPC_ gene and carbapenemase-producing Citrobacter were almost undetectable in patients during the study period: only one patient carried a VIM-producing *C. freundii*. These discrepancies could be due to a specific resident community in the wastewater network, or to our limited knowledge of KPC-producing bacteria and Citrobacter spp. in the current CPE epidemiology. The high prevalence of carbapenemase-producing *C. freundii* and *C. braakii* in wastewater is concerning, given that these bacteria are increasingly being reported in healthcare-associated infections [Yao et al., 2021, Fonton et al., 2024]. Moreover, evidence that Citrobacter species are emerging carriers of carbapenem-resistance genes has recently been published [Wang et al., 2025]. The role of environmental reservoirs (U-bends, sinks, etc) of carbapenemase-producing *C. freundii* in persistent outbreaks of CPE was demonstrated [, Fonton et al., 2024]. Since our study, the regional epidemiology changed with more and more carbapenemase-producing Citrobacter described in hospital outbreak (ICT surveillance data 2023-2024). A recent study in Germany showed that *C. freundii* is the most common species producing carbapenemases in humans at the national level [Sommer et al., 2024].

The abundance of carbapenemase-producing Citrobacter spp., Aeromonas spp. and Pseudomonas spp. in the microbial community of effluent network suggested a possible exchange of emerging carbapenemase genes between human and environmental bacteria. Indeed, Citrobacter spp., Aeromonas spp. and Pseudomonas spp. can also colonise the human microbiota and act as a shuttle between humans and the environment. Aeromonas spp. were recently described for the first time as carriers of *bla*_OXA-48_ and *bla*_KPC_ [Drk et al., 2023]. This led to the hypothesis of an exchange of ARG from human bacteria to autochthonous wastewater bacterial communities, with some species and resistotypes finding favourable conditions in the wastewater network. Therefore, wastewater could be used as an indicator of the further emergence of AMR and predict human risk, even if the signals are not related to real-time clinical epidemiology.

The second objective of this study was to compare wastewater carbapenemase genes between two different patient populations characterised by different risks for CPE and VRE carriage or infection. Significant differences were observed between the HRR and SRR in the load of AMR genes and the rate of cultivable CPE isolates. For the majority of genes and resistotypes, a higher prevalence was observed in the HRR, corresponding to the increased risk of CPE and VRE carriage in HRR patients. Majlander et al (2021) have previously observed this between two hospitals in Finland. This suggests that despite the low correlation between the wastewater AMR signal and individual patient cases on any given day, the quantification of carbapenemase genes in wastewater may be able to detect differences and thus monitor changes in epidemiology at the population level. In this later indication, WWS of AMR could be a valuable and efficient tool for local infection control.

## Conclusion

In summary, we have demonstrated that the assessment of emerging AMR in wastewater can be carried out in a real hospital environment, subject to adjustments to the sampling programme based on patient flow and network structure. Even if WWS distinguished two contrasting populations of patients in our hospital, the WWS signals were only partially consistent with local epidemiology, suggesting that interpreting the WWS of AMR will be more challenging than interpreting the WWS of viruses. This is probably due to the fact that bacteria can colonise wastewater networks, multiply and exchange genes. The results support that local AMR WWS for IC purposes should be prospective and continue. This is necessary for modelling ARG dynamics, setting threshold alerts, linking WWS signals to infection control policies, and evaluating WWS in predictive epidemiology.

## Appendices

**Supplementary Table S1.**
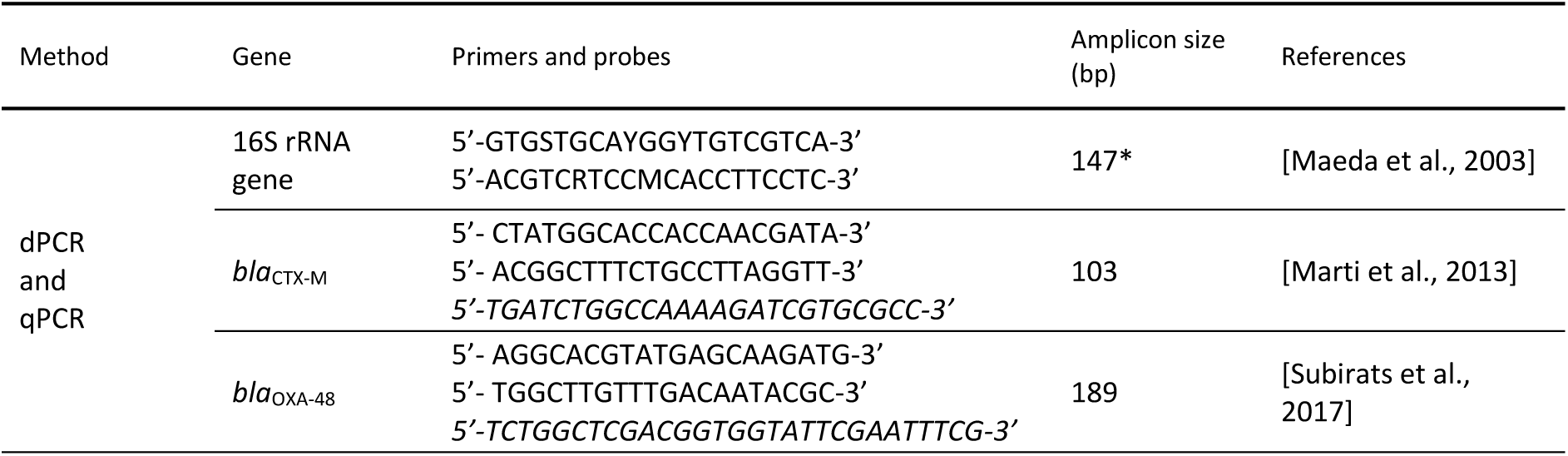
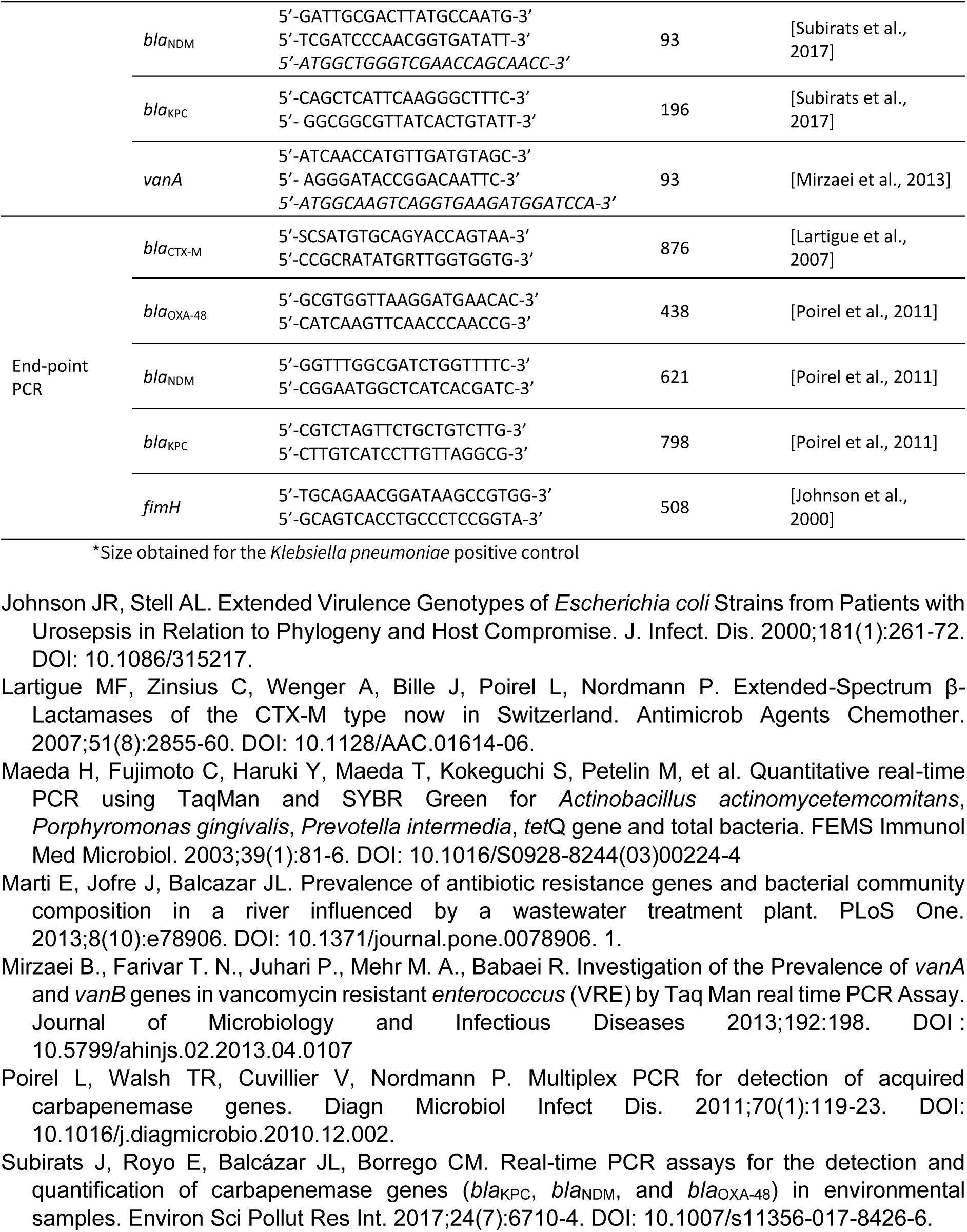
Primers and probes used for qPCR, dPCR and end-point PCR. Probes for dPCR are indicated in italics.

**Supplementary Table S2.** Characteristics of standard qPCR ranges.

| Amplified gene | Specific Tm of the standard range | Last quantifiable range point<br>(gene copy number/reaction) |
| --- | --- | --- |
| 16S rRNA gene | 85.94 ± 0.05 °C | 57.0 |
| *bla<sub>CTX-M</sub>* | 85.36 ± 0,01 °C | 4.8 |
| *bla<sub>OXA-48</sub>* | 83.90 ± 0.05 °C | 5.7 |
| *bla<sub>NDM</sub>* | 90.16 ± 0.06 °C | 5.0 |
| *bla<sub>KPC</sub>* | 88.71 ± 0.1 °C | 6.2 |
| *vanA* | 79.82 ± 0.1 °C | 5.8 |

**Supplementary Figure S1.**
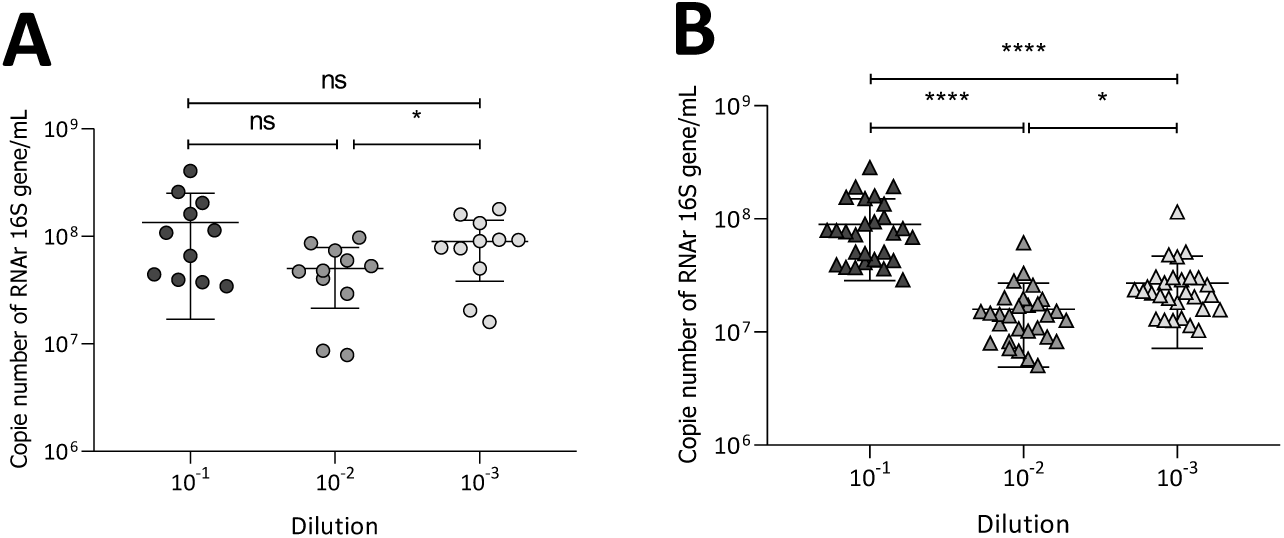
Distribution of concentrations of 16S rRNA gene in SRR (A) and HRR (B) wastewaters. The results of environmental DNA qPCR (A) with 10^-1^, 10^-2^ and 10^-3^ dilutions are expressed on a logarithmic scale. Comparison of quantification between dilutions was performed by the unpaired t test or the Mann-Whitney test, and p-values are presented above the box plots (ns: non-significant; *0.01 < p-value ≤ 0.05; ***0.0001 < p-value ≤ 0.001; ****p-value ≤ 0.0001). Median and IQR are presented on each scatter plot. For HRR samples, Pearson correlation coefficients indicated a strong positive correlation between dilutions (r ≥ 0.71, p < 1.58E-05) suggesting consistent quantification across dilutions. For SRR samples, Spearman correlation coefficients were also high (p ≥ 0.59), but with slightly greater variability, particularly between 10⁻¹ and 10⁻³ (p = 0.59, p = 0.0609).

## Acknowledgements

We thank the PHySE-HSM team, Sébastien Mercier from the hospital plumbing workshop, and the Hospital Infection Control team at Montpellier Hospital for their help in collecting samples and data and processing the results. The authors thank Philippe Clair, director of the qPCR facility at the University of Montpellier (Montpellier GenomiX), for his technical advice and for the loan of a LightCycler Nano during the study. The authors also thank Elodie Pichon for excellent dPCR technical assistance.

## Data, scripts, code, and supplementary information availability

The datasets used and/or analysed during the current study are available from the corresponding author on reasonable request.

